# The potential impact of intervention strategies on COVID-19 transmission in Malawi: A mathematical modelling study

**DOI:** 10.1101/2020.10.06.20207878

**Authors:** Tara D. Mangal, Charlie Whittaker, Dominic Nkhoma, Wingston Ng’ambi, Oliver J Watson, Patrick Walker, Azra Ghani, Paul Revill, Tim Colbourn, Andrew Phillips, Timothy B. Hallett, Joseph Mfutso-Bengo

## Abstract

**Background:** COVID-19 mitigation strategies have been challenging to implement in resource-limited settings such as Malawi due to the potential for widespread disruption to social and economic well-being. Here we estimate the clinical severity of COVID-19 in Malawi, quantifying the potential impact of intervention strategies and increases in health system capacity.

**Methods:** The infection fatality ratios (IFR) in Malawi were estimated by adjusting reported IFR for China accounting for demography, the current prevalence of comorbidities and health system capacity. These estimates were input into an age-structured deterministic model, which simulated the epidemic trajectory with non-pharmaceutical interventions. The impact of a novel therapeutic agent and increases in hospital capacity and oxygen availability were explored, given different assumptions on mortality rates.

**Findings:** The estimated age-specific IFR in Malawi are higher than those reported for China, however the younger average age of the population results in a slightly lower population-weighted IFR (0.48%, 95% uncertainty interval [UI] 0.30% – 0.72% compared with 0.60%, 95% CI 0.4% – 1.3% in China). The current interventions implemented, (i.e. social distancing, workplace closures and public transport restrictions) could potentially avert 3,100 deaths (95% UI 1,500 – 4,500) over the course of the epidemic. Enhanced shielding of people aged ≥ 60 years could avert a further 30,500 deaths (95% UI 17,500 – 45,600) and halve ICU admissions at the peak of the outbreak. Coverage of face coverings of 60% under the assumption of 50% efficacy could be sufficient to control the epidemic. A novel therapeutic agent, which reduces mortality by 0.65 and 0.8 for severe and critical cases respectively, in combination with increasing hospital capacity could reduce projected mortality to 2.55 deaths per 1,000 population (95% UI 1.58 – 3.84).

**Conclusion:** The risks due to COVID-19 vary across settings and are influenced by age, underlying health and health system capacity.

**Summary Box:** *What is already known?:*

- As COVID-19 spreads throughout Sub-Saharan Africa, countries are under increasing pressure to protect the most vulnerable by suppressing spread through, for example, stringent social distancing measures or shielding of those at highest risk away from the general population.
- There are a number of studies estimating infection fatality ratio due to COVID-19 but none use data from African settings. The estimated IFR varies across settings ranging between 0.28-0.99%, with higher values estimated for Europe (0.77%, 95% CI 0.55 – 0.99%) compared with Asia (0.46%, 95% CI 0.38 – 0.55).
- The IFR for African settings are still unknown, although several studies have highlighted the potential for increased mortality due to comorbidities such as HIV, TB and malaria.
- There are a small number of studies looking at the impact of non-pharmaceutical interventions in Africa, particularly South Africa, but none to date have combined this with country-specific estimates of IFR adjusted for comorbidity prevalence and with consideration to the prevailing health system constraints and the impact of these constraints on mortality rates.

*What are the new findings?:*

- After accounting for the health system constraints and differing prevalences of underlying comorbidities, the estimated infection fatality ratio (IFR) for Malawi (0.48%, 95% uncertainty interval 0.30% – 0.72%) is within the ranges reported for the Americas, Asia and Europe (overall IFR 0.70, 95% CI 0.57 – 0.82, range 0.28 – 0.89).
- Introducing enhanced shielding of people aged ≥ 60 years could avert up to 30,500 deaths (95% UI 17,500 – 45,600) and significantly reduce demand on ICU admissions.
- Maintaining coverage of face coverings at 60%, under the assumption of 50% efficacy, could be sufficient to control the epidemic.
- Combining the introduction of a novel therapeutic agent with increases in hospital capacity could reduce projected mortality to 2.55 deaths per 1,000 population (95% UI 1.58 – 3.84).

*What do the new findings imply?:*

- Adjusting estimates of COVID-19 severity to account for underlying health is crucial for predicting health system demands.
- A multi-pronged approach to controlling transmission, including face coverings, increasing hospital capacity and using new therapeutic agents could significantly reduce deaths to COVID-19, but is not as effective as a theoretical long-lasting lockdown.

## INTRODUCTION

As of 22^nd^ August 2020, the novel coronavirus SARS-CoV-2 had spread throughout 216 countries, with over 21 million cases and 760,000 deaths worldwide.^1^ In response to this threat, numerous African countries acted swiftly, implementing a suite of control measures aimed at slowing down the spread of disease and minimising deaths. On 20^th^ March 2020, Malawi declared a state of national disaster, ordering all schools to close, barring large gatherings and restricting international travel. This was followed by the order of a 21-day lockdown to begin on 18^th^ April 2020, which was later suspended by the High Court of Malawi due to concerns around the implications on vulnerable populations. Further control measures were introduced in April, including social distancing and restrictions on public transport use. The first three cases in Malawi were confirmed on 2^nd^ April 2020 and by 8^th^ July 2020, clusters of cases had occurred across 28 districts (1,818 confirmed cases with 19 deaths) although widespread community transmission had not been reported.^1,2^

Given the limited capacity of the health system in Malawi to respond to an emerging outbreak and the likelihood that effective pharmaceutical interventions may not be available for many months, there is a critical reliance on non-pharmaceutical interventions (NPI) to reduce transmission. These measures include isolation of suspected/confirmed cases, contact tracing, social distancing, travel restrictions, face covering, school and workplace closures and shielding of the most vulnerable.^3-6^

The impact of NPI can be summarised as a change in the effective reproduction number *Rt*, which represents the average number of secondary infections resulting from one infected case. Strict interventions such as lockdown, where population movement is limited to only essential travel and most public facilities and transport links are closed, have shown the most success in reducing transmission.^5,7^ However major restrictions to working practices or public transport may have catastrophic implications in Sub-Saharan Africa, where many have limited financial capacity to withstand income shocks and no access to social protection programmes.^8^ Face coverings have been recommended by WHO as one possible intervention which could reduce transmission of SARS-CoV-2 with minimal socioeconomic implications despite a lack of high-quality evidence.^9,10^ Nevertheless, several countries, (including, Canada, South Korea and Czech Republic), have made face coverings mandatory in public spaces and the UK has made the use of face masks mandatory on public transport and in shops.

The impact of NPI on SARS-CoV-2 transmission in Malawi depends critically on the local context such as population behaviour (including uptake of and compliance with such measures), population movement and contact patterns (Malawi is over 80% rural and many rely on subsistence farming) and health system capacity.^11^

A key priority during this emerging pandemic, is estimating clinical severity and health system requirements. Current oxygen capacity in hospitals may not be sufficient to give supportive care to large numbers of severe COVID-19 cases; 65% of 34 hospital wards in Malawian hospitals recently assessed had (what was considered pre-COVID-19) adequate access to oxygen and priority is now being given to scaling up access and supplies urgently.^12,13^ The impact of oxygen supplementation on COVID-19 mortality is not well established however early studies suggest oxygen could reduce the need for mechanical ventilation and lower the risk of death.^14^ Several therapeutic agents have been considered for treatment of severe COVID-19, two of which, Dexamethasone and Remdesivir, have been effective in improving patient outcomes.^15,16^

Infection fatality ratios (IFR), defined as the number of deaths divided by the number of infections are challenging to estimate, particularly in an emerging outbreak, due to the difficulties in identifying the true number of infected people (both asymptomatic and symptomatic). IFR are strongly dependent on age and the majority of deaths reported early in the epidemic were among those aged over 60 years.^17-19^ There is also now a growing body of evidence on the elevated risk of mortality with certain underlying comorbidities, such as cardiovascular disease (CVD), diabetes, chronic obstructive pulmonary disease (COPD) and infectious diseases, for example HIV, TB and malaria.^20-23^ The majority of these data were reported from high-income settings, which have borne the highest burden of disease recorded so far. It is not yet clear how these risk factors will affect disease severity in countries like Malawi, which have a younger population overall, but a high prevalence of infectious diseases and untreated chronic conditions.

### Objectives

Three objectives form the focus of this paper: 1) estimate disease severity caused by SARS-CoV-2 in the Malawian population given its demographic structure, the prevalence of key comorbidities and health system capacity; 2) examine the potential impact of a range of NPI that have been or could be used in Malawi; 3) on that basis, investigate the potential extent to which increasing health system capacity and/or providing therapeutics could contribute to reducing deaths due to SARS-CoV-2 infection.

## METHODS

We present the methods in three sections that relate to each of our aims.

### 1. Estimates of Infection Fatality Ratios in Malawi

Our approach utilises data on age-specific IFR from China (one of the few studies which applies demography-adjusted under-ascertainment corrections) and then makes adjustments based on the relative burdens of diseases relevant to COVID-19 risk between China and Malawi, making assumptions about the extent to which each disease affects IFR.^19^

The prevalence of HIV (virally suppressed and unsuppressed), active TB, clinical malaria, CVD, COPD, hypertension, diabetes (type I and II), obesity (defined as BMI ≥28 kg/m^2^ according to Chinese criteria and ≥30 kg/m^2^ using Malawian criteria) and malnutrition were extracted for Chinese and Malawian populations (see Supplementary Table 1 for data sources). We created a unified risk factor for “metabolic syndrome”, the prevalence for which was the greatest of the prevalences of CVD, hypertension, obesity and diabetes due to clustering of these conditions. The risk for metabolic syndrome was taken as the highest risk reported for any of the pooled conditions. Given the considerable uncertainty in these estimates along with likely differences across settings, we considered a wide range of relative risk values for each comorbidity (Supplementary Table 2).

The age-distributed estimates for IFR in Malawi were derived from those published by Verity *et al* for cases reported in mainland China, using linear interpolation on the log scale to derive values in five-year age-groups from the 10-year age-bands reported.^19^ Adjusted IFR estimates by age for Malawi were computed as follows:

1. Lognormal distributions were derived for each of the age-distributed IFR from China such that the mean matched the mean IFR and 95% of the probability mass fell inside the reported 95% bounds. Uniform distributions were defined for the relative risks of mortality due to COVID-19 for each comorbidity covering the range described in Supplementary Table 2
2. Age-specific IFR and relative risks were sampled from the defined distributions
3. Age-distributed IFR for a theoretical population with no comorbidities (*IFR\**_*a*_) were computed using Equation 1:

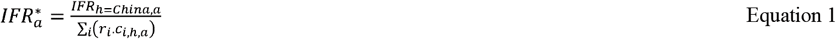 Where *IFR*_*h=China,a*_ is the sampled IFR in setting *h* (where *h* is China), *i* is the index for each comorbidity, *r*_*i*_ is the sampled relative risk of mortality for each condition and *c*_*i,h,a*_ is the prevalence of each comorbidity in setting *h*. All terms except relative risk values are additionally indexed by age-group *a*.
4. The adjusted IFR for Malawi were then estimated using Equation 2:

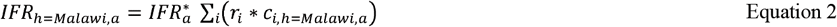
5. Steps 2-4 were repeated 1000 times and the median adjusted IFR and uncertainty intervals were calculated as the 50^th^, 2.5^th^ and 97.5^th^ quantiles from the sampled estimates

A summary for the average IFR for Malawi was obtained by weighting the age-specific IFR by the proportion of the population in each age-group, as follows:

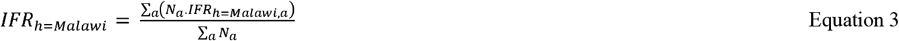

Where *N*_*a*_ is the number of persons in that age-group.

This method assumed that the differences between IFR in Malawi compared with China are due to comorbidities and age-structure: the effect of access to and quality of healthcare was accounted for through the following additional steps:

1. Parameter sets defining the proportion of COVID-19 cases requiring different levels of hospital care (severe or critical) were generated using rejection sampling on the basis of (i) prior information from high-income settings; (ii) assumptions for the mortality according to disease severity with treatment; and, (iii) accepting those parameter sets which resulted in an IFR which fell within the IFR computed for this setting under the assumption of similar healthcare capacity.
2. These estimated proportions of disease severity were used in the simulation model, with additional assumptions about mortality in the absence of care (severe cases experience an age-dependent mortality rate which is doubled if no treatment is received; mortality rates are 0.5 for critical cases receiving treatment and 0.95 if untreated).
3. Epidemic trajectories were simulated in which the availability of hospital care is limited to the level currently prevailing in Malawi. The induced overall IFR in Malawi given the current healthcare system constraints at the end of the epidemic is “Number died of COVID-19 / Number ever infected with SARS-CoV-2”.

Further details are provided in the SI.

### 2. Estimates of the Potential Impact of NPI on Transmission of SARS-CoV-2

The COVID-19 Model of Walker *et al*. was used to make projections of the spread of SARS-CoV-2 in Malawi under a number of NPI scenarios.^24^ Briefly, the model comprises an age-structured compartmental deterministic framework which describes the transmission of SARS-CoV-2 through an otherwise homogenous population. The rates of contact between age-groups are derived from the Manicaland study in Zimbabwe (Supplementary Table 4).^25^ Infected (and infectious) cases are classified as mild (not requiring care), severe (requiring hospitalisation / oxygen) and critical (requiring ICU / mechanical ventilation) with the likelihood of receiving care constrained by the prevailing health system capacity. We assume that not all critical cases will require mechanical ventilation, but for those that do, access to an ICU bed will also indicate availability of mechanical ventilation. In each infected stage, there is a probability of death, derived from multinational analyses from data in China, UK and US. It is assumed that hospitalised cases will not contribute to transmission. In the case of a person needing care but health system capacity being exhausted, the person is exposed to a risk of death consistent with no care being received (Supplementary Table 3). The probability of death in severe cases not receiving treatment is lowered from the default values in the Walker *et al* model to double that of treated cases to reflect the low mortality observed in Malawi to date.^26^

We assumed *R*_*0*_ = 2, which is a central value for the estimates of *R*_*0*_ in Malawi before nationwide non-pharmaceutical interventions were introduced and vary this between 1.5 and 3 to reflect the uncertainty in this assumption.^27^ At the start of the outbreak, 20 cases were seeded in age-groups 35-54 reflecting the ages of those who are most likely to have travelled internationally and acquired infection.

We ran each simulation 1000 times over 250 modelled days (365 days when changing the duration of NPI) using the sampled parameter sets for disease severity (see above). The outcomes of each intervention on the daily numbers infected, health system requirements (broken down by severity) and deaths are presented.

#### Non-pharmaceutical intervention strategies

The interventions under consideration are shown in Supplementary Table 4. The duration of lockdown was varied between 6 and 24 weeks, whereupon the previously implemented intervention strategies were resumed.

Face coverings were analysed as an incremental intervention on top of the existing measures and we explored a full range of values for efficacy and proper usage assuming that the current measures in place remain for the duration of the simulation. We don’t distinguish between household and non-household transmission and assume adherence and efficacy jointly reduce the risk of transmission to the whole population.

The low numbers of deaths reported to date in Malawi coupled with the high potential for under-reporting make formal calibration to surveillance data problematic. We opt, therefore, to present a hypothetical future scenario where community transmission has started to occur.

### 3. Estimates of the impact of increasing health system capacity

The projected effects of increasing the number of non-intensive care hospital beds plus availability of oxygen and the introduction of a novel therapeutic agent were examined assuming that the current intervention strategies would remain in place indefinitely. We simulated an increase in hospital bed capacity (plus access to oxygen) by up to 100% from the trigger day of 1.0 deaths per 100,000 population, presenting the resulting impact on the cumulative number of deaths projected to occur over the epidemic, which depends critically on the assumptions around mortality in severe cases. We conservatively estimated age-dependent mortality rates would double if severe cases were untreated but given the uncertainty in this, explored an additional scenario in which the untreated mortality rate was 0.6 for all age-groups.^24^ Additionally, the impact of a novel therapeutic agent was analysed, assuming a proportional reduction in mortality for severe and critical cases (0.65 and 0.8 respectively, based on the findings from the RECOVERY trial on Dexamethasone) applicable to all age-groups. We assumed that the therapeutic agent could be administered to those in need even if hospital beds or ICU beds were not available.

All analyses were conducted in R statistical software, version 3.6.3 (https://www.r-project.org/). Source code and supporting documentation for the SEIR model is available at https://github.com/mrc-ide/squire.

### Patient and Public Involvement

Patients and/or the public were not involved in the design, or conduct, or reporting, or dissemination plans of this research.

### Role of the funding source

The funders of this study had no role in study design, data analysis, data interpretation or writing of the report. All authors had access to all data in the study and accept final responsibility for the decision to submit for publication.

## RESULTS

### 1. Estimates of the infection fatality rate in Malawi

The estimated age-specific IFR for Malawi are higher for every age-group than those reported in China under the assumption of similar healthcare availability (Figure 1). However, the estimated population-weighted IFR are lower in Malawi (IFR 0.24%, 95% uncertainty interval [UI] 0.12 – 0.61%, compared with 0.6%, 95% confidence interval 0.40% – 1.30% in China) due to the younger average age of the population. When incorporating the known health system capacity constraints in Malawi through the simulation model, overall population-weighted IFR increase but remain slightly lower than the estimates for China although the difference is not significant (overall IFR 0.48%, 95% UI 0.30% – 0.72%).

**Figure 1.**
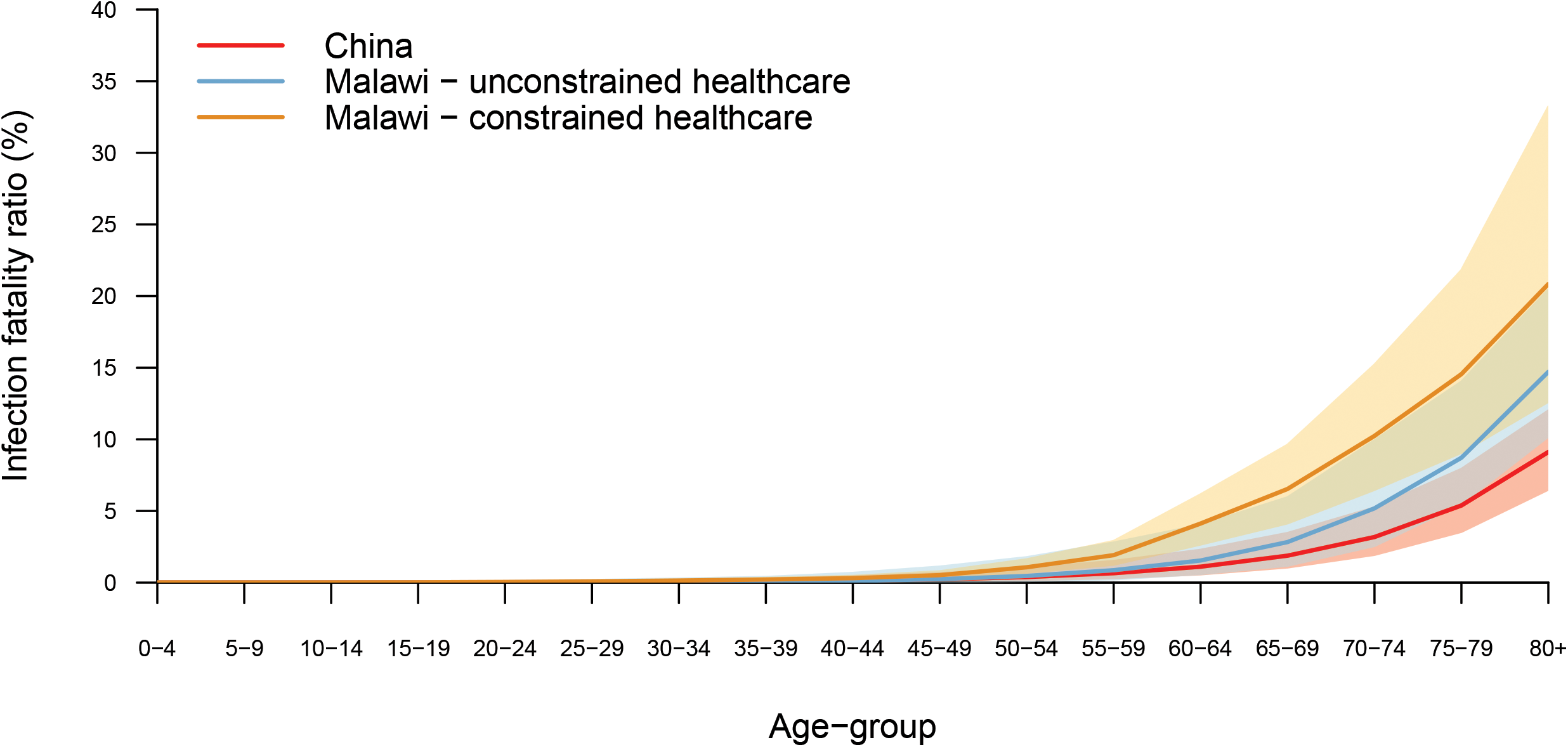
Adjusted estimates of infection fatality ratios for the Malawian population with unconstrained healthcare and constrained healthcare (according to current health system capacity) compared with estimates from China.

### 2. Estimates of the potential impact of NPI on transmission of SARS-CoV-2

The projected unmitigated scenario is presented as a counterfactual, showing what could occur had no interventions been introduced (Figure 2). With the current interventions in place, and assumed to be in place indefinitely, we estimate approximately 3,100 deaths (95% UI 1,500 – 4,500) could be averted over the course of the epidemic compared with an unmitigated scenario in which 80,400 deaths (95% UI 49,500 – 118,700) are projected to occur (Table 1). The projected death rate over the 250 modelled days with the current interventions in place is 4.20 deaths per 1,000 population (95% UI 2.51-6.00 deaths per 1,000 population). These measures would also delay the spread of infection, shifting the peak in infections by approximately 11 days.

**Table 1.** Outputs from intervention strategies over 250 days

|  | Unmitigated | Current situation | Enhanced shielding | Lockdown |
| --- | --- | --- | --- | --- |
| Total cases / 1,000 | 770<br>(666-875) | 584<br>(507-662) | 513<br>(436-595) | 71<br>(58-85) |
| Number hospital beds required at peak | 40,000<br>(27,800-59,900) | 35,600<br>(25,300-53,100) | 27,900<br>(17,800-38,000) | 2,100<br>(1,100-3,500) |
| Number ICU beds required at peak | 2,100<br>(1,400-3,400) | 1,800<br>(1,200-2,900) | 900<br>(600-1,400) | 100<br>(100-200) |
| Total deaths / 1,000 | 4.2<br>(2.6-6.2) | 4.0<br>(2.5-6.0) | 2.4<br>(1.63-64) | 0.5<br>(0.3-0.8) |

All values are medians of 1000 simulations using the sampled parameter sets for disease severity. Median hospital and ICU beds are rounded to the nearest 100.

**Figure 2.**
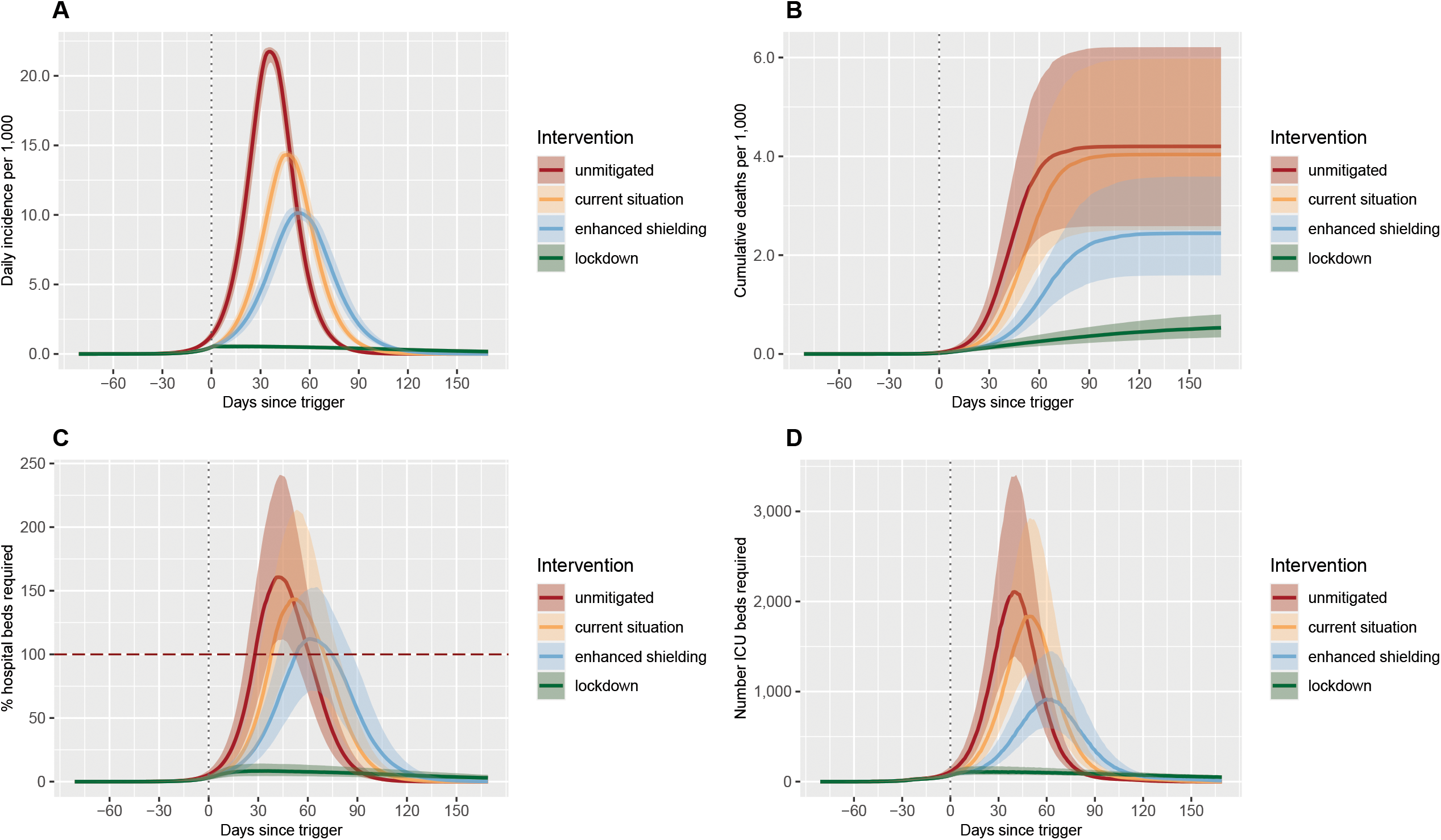
Impact of non-pharmaceutical interventions compared with a baseline (unmitigated) scenario on the daily incidence per 1,000 population (A), the cumulative deaths per 1,000 population (B), the percentage of hospital beds that are required (C) and the number of ICU beds that are required (D). The unmitigated scenario presents the counterfactual situation had no interventions been introduced. The current situation reflects the non-pharmaceutical interventions adopted by Malawi at the beginning of the outbreak. Enhanced shielding refers to reducing contact rates of people aged ≥ 60 years. Lockdown is the adoption of stringent social distancing policies. Further details are presented in Table 1. The trigger date is shown with a vertical grey dashed line. The red horizontal dashed line shows the capacity of the health system for non-intensive care (C). ICU capacity comprises 25 ICU beds and 16 mechanical ventilators.^33^

The predicted age distribution of infected people at the peak of the epidemic shows that the majority of infections occur in the younger ages (<20 years) which make up >50% of the population and have high contact rates (Supplementary Figure 1), although only 1.3% of deaths occur in that group. There is, however, considerable uncertainty around the prevalence and impact of comorbidities such as HIV and malnutrition in these age-groups. The majority of projected deaths occur in those aged over 70 years.

Of mitigation strategies modelled, long-term lockdown has the largest impact, bringing the infection rate down to 71 infections per 1,000 population (95% UI 58– 85 per 1,000 population) and the mortality rate to 0.5 deaths per 1,000 population (95% UI 0.3-0.8 per 1,000 population, equivalent to 10,100 deaths, 95% UI 6,400 – 15,300, Table 1). Applying lockdown over 6, 12 and 24 weeks delays the peak incidence but an outbreak with a similar magnitude to the projections under the current situation would still occur after lockdown is relaxed in the absence of any further interventions (Supplementary Figure 2).

With current interventions in place, coverage of face coverings would need to exceed 60% (50% with shielding implemented simultaneously) with a minimum of 50% efficacy in order to reduce projected *Rt* to below 1 (Figure 3).

**Figure 3.**
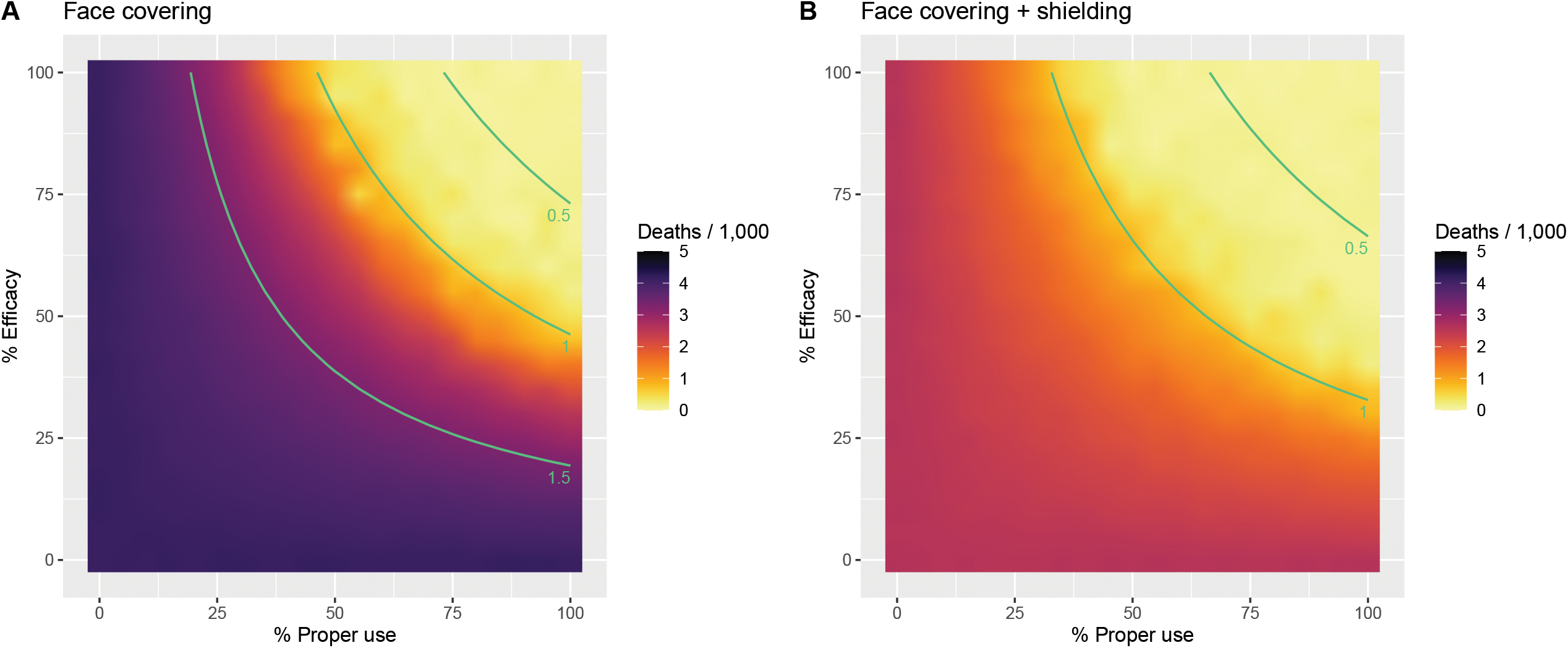
Impact of face covering on the total number of deaths per 1,000 population projected to occur over 250 days. The isoclines (green lines) represent the estimated *Rt* given the percentage efficacy and percentage of proper use (adherence). The impact of face covering (and enhanced shielding [B]) is assumed to be incremental to the current interventions in place.

### 3. Estimates of the impact of increasing health system capacity

If it is assumed that mortality is low in severe cases, then increasing capacity for non-intensive care (hospital beds with oxygen availability) by 50% reduces the projected mortality rate to 3.89 deaths per 1,000 population (95% UI 2.47 – 5.79) compared with 4.04 (95% UI 2.51 – 5.98) under the current scenario (Supplementary Table 8). Doubling hospital and oxygen capacity could only marginally further reduce this to 3.79 deaths per 1,000 population (95% UI 2.45 – 5.61). If, instead, we assume that the probability of death in untreated severe cases is 0.6 across all ages (consistent with the clinical estimates used by Walker *et al* derived from high-income settings), the proportional reduction in expected deaths would be larger, falling from 5.96 deaths per 1,000 population (95% UI 2.99 – 10.62) in the current scenario to 4.41 deaths per 1,000 population (95% UI 2.48 – 7.87) with double the hospital bed capacity.^24^ Introducing a novel therapeutic agent that is capable of reducing mortality by 0.6 for severe cases by 0.85 for critical cases, in combination with a 50% scale up in hospital capacity reduces projected deaths to 2.55 deaths per 1,000 population (95% UI 1.58 – 3.84, Figure 4).

**Figure 4.**
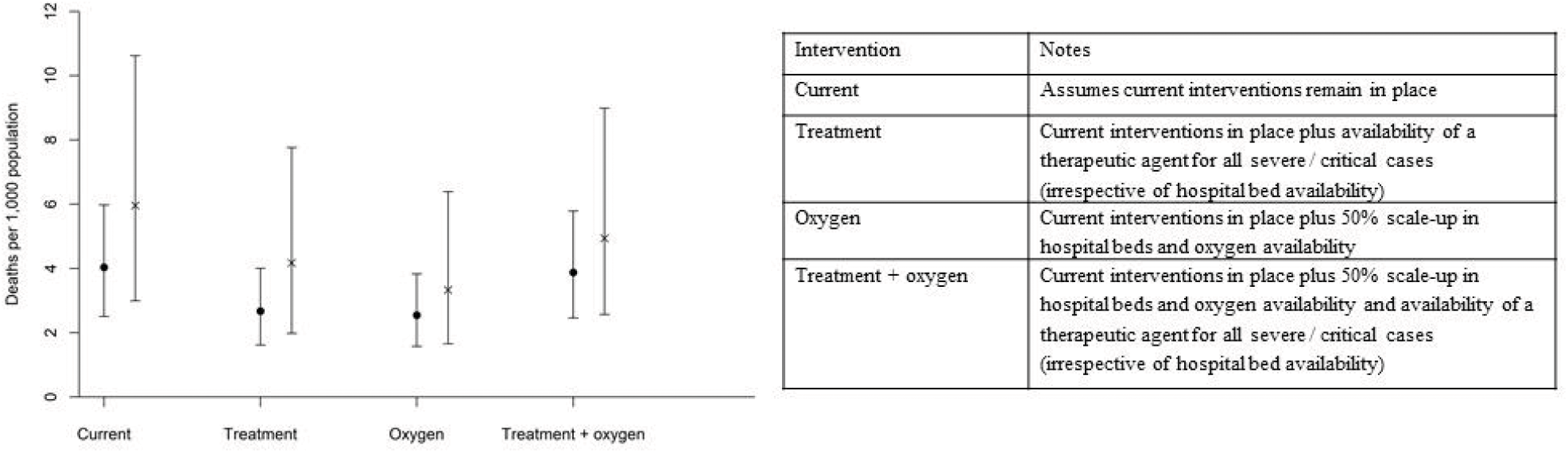
Projected total numbers of deaths per 1,000 people over 250 days with increases in hospital capacity (and oxygen) and a novel therapeutic agent.

## DISCUSSION

The results shown here give several important insights into the potential spread and severity of SARS-CoV-2 infection in Malawi and what can be done to prepare for it.

First, under the assumption of similar access to healthcare, estimated age-specific IFR in Malawi are higher compared with China although overall population-weighted IFR are lower due to the younger average age of the population.^19^ As the availability of health care is much lower in Malawi, the overall expectation for IFR increases to 0.48% (95% UI 0.30% – 0.72%), slightly lower although still within the range of estimates reported across the Americas, Asia and Europe (overall IFR 0.70, 95% CI 0.57 – 0.82, range 0.28 – 0.89).^28^ Estimates of mortality in Uganda follow a similar trend, with a lower predicted disease burden than the comparator regions (Europe, America and China) and the majority of risk arising indirectly due to disruptions to the health service.^29^ Other studies have also reported a differential risk of severe disease in African settings compared with European countries due to underlying health conditions although there is considerable uncertainty in these analyses.^30,31^

Second, we find that the intervention strategies that have been implemented so far in Malawi would be unlikely to suppress or substantially mitigate the epidemic. Although lockdowns have been highly effective across a number of settings, it is may be impractical in Malawi for a number of reasons.^5,32,33^ Approximately 50% of the Malawian population lives in poverty, meaning there is no financial buffer if people are unable to earn money.^34^ The disruption to food production and delivery chains is likely to impact those who are most vulnerable, with food shortages likely to occur within days of lockdown being implemented.

In principle, shielding of the elderly (plus other vulnerable populations, such as people living with HIV or TB) should be effective in significantly reducing the death rate. The practicalities of moving elderly people into separate accommodation in the African context are uncertain at best but it is a potentially risky strategy as a single imported case could have devastating effects. In high-income countries, this strategy has proven difficult with outbreaks occurring in many nursing homes.^35^ Designating shielded households, or even single rooms within a household may be a viable strategy in Malawi which has an extensive network of community health workers who could facilitate and support such measures.

Given the difficult choices facing decision-makers in Malawi and elsewhere, it is not surprising that significant attention has turned to less disruptive strategies for restricting some population movement and advocating use of face coverings, a putatively low-cost and readily available means of reducing transmission risk. However, there is no clear indication as yet whether widespread proper use of face coverings would be possible, and thus it remains highly uncertain how effective face covering will be in reducing risk of SARS-CoV-2 transmission. Delaying the epidemic by even a few months could allow time for new therapeutic interventions to be implemented subject to evidence of effect.^36^ Additional interventions, such as contact tracing and testing (test, trace, isolate) and local quarantining could have a significant impact on the spread of infection and have been regularly used for case-finding and containment for HIV and TB in low and middle-income countries.^37-40^ However they are extremely intensive and difficult to implement at a large scale and therefore may not be a feasible option for mitigating the outbreak in Malawi.

Third, we find that increases in certain types of capacity in hospital may contribute to reducing deaths. The current priorities for COVID-19 response in Malawi are to significantly expand access to oxygen concentrators in existing hospitals through the acquisition of new concentrators or using splitters to increase capacity.^13^ The expected impact of this is driven by the assumptions on the mortality of severe cases, for which very few data from low-income countries are available to inform. The potential benefit is largest when the estimated mortality in severe cases is high. Additionally, there are likely to be improvements to long-term morbidity and lung health if severe cases receive oxygen when required, combined with reduced probability of requiring mechanical ventilation, although we do not capture this here. We define individuals as mild, severe or critical from the outset and do not include a probability of progression from severe to critical, which could be reduced by hospital bed and oxygen availability. There is an urgent need for further data to analyse the longer-term impacts of COVID-19 infection to inform this. Randomised trials of treatments for COVID-19 are underway in many countries and the introduction of a low-cost effective therapeutic agent, such as Dexamethasone or (generic) Remdesivir, could have the potential to significantly reduce clinical severity.^15,16,36^

The IFR that we use as our baseline along with some key parameters including treatment outcomes rely on data from mainland China, North America and Europe which may not be directly transferable to Malawi.^5,19,24^ Our results are sensitive to the assumptions inherent in these analyses and cannot yet be fully parameterised using data from African settings due to the limited numbers of cases there. Age-structured contact matrices are derived from studies in Zimbabwe although we expect that there are unlikely to be significant differences between the two settings that would meaningfully affect the conclusions drawn. Household structure is not incorporated therefore we cannot adjust for increased risk of infection within households. This may be of particular importance in Malawi where households are multigenerational and there may not be space to designate separate rooms for high-risk individuals. The absence of nosocomial transmission in this model may result in an under-estimation of the incidence of SARS-CoV-2 if PPE availability within hospitals is sub-optimal. An important next step in this analysis would be the integration of the model with geographically-disaggregated surveillance data on testing, deaths, movement and other data which could capture local transmission and potentially open the way to more finely targeted interventions that may maximise epidemic control with lesser disruptions overall.

The estimates of relative risk of mortality with comorbidities are derived mainly from studies in high-income settings and only one is based on data from sub-Saharan Africa. These risks may vary by age although we do not capture this here. The management of comorbidities is likely to differ across settings and so the corresponding risk of mortality with these conditions may vary also. Lastly, we do not distinguish between oxygen requirements or mechanical ventilation needs of severe and critical cases although the mortality benefits from a novel therapeutic agent could vary between these groups.

The slow initial spread of SARS-CoV-2 up to 20^th^ June 2020 could be consistent with a lower *R*_*0*_ than is assumed here, an imperfect surveillance system with low numbers of tests being carried out, or few introductions of cases to Malawi with measures having been sufficient so far in limiting transmission.^27^ This study has focussed on the effects of non-pharmaceutical interventions and scaling up health system capacity in respect of one disease, COVID-19. However imposing lockdown could disrupt routine health services such as the provision of care for HIV, tuberculosis and malaria along with national immunisation programmes, compounding increases in mortality rates.^41^ Balancing the competing demands on health versus economic productivity, poverty and education is an extremely difficult decision and we present these projections as a series of hypothetical scenarios which could be used to inform decision-making.

These outputs are not intended to be a forecast of what will happen in Malawi and evidence should be reviewed in the light of the continuously evolving surveillance data and combined with detailed analyses on the broader impacts of any potential interventions. Lessons could be learnt from the South African response, which implemented a rapid, phased strategy, successfully delaying the outbreak despite considerable challenges. In addition to physical distancing and restrictions on movement, South Africa capitalised on existing experienced teams of community health workers to conduct active case-finding along with redirecting contact-tracing teams, previously established for TB control, to conduct COVID-19 contact-tracing and monitor quarantine compliance.^38^ Clearly lockdown would have the biggest impact on spread of SARS-CoV-2, however in settings where this is not feasible, a combination of interventions such as shielding, face covering, increasing hospital capacity and therapeutic agents could together have a significant impact on mortality.

## Data Availability

All data used in the analyses are publicly available and sources have been listed for each.

https://github.com/mrc-ide/squire

## Contributors

Literature search: TDM, TC, AP, TBH

Figures: TDM

Study design: All authors

Data collection: TDM, CW, DN, WN, OJW, PW, AG, TBH, JM-B

Data analysis: TDM, TBH, AP, TC

Data interpretation: All authors

Writing: All authors

## Declaration of interests

The authors declare no conflicts of interest.

## Acknowledgments

Dr Mangal, Prof Hallett, Prof Phillips, Dr Colbourn, Prof Mfutso-Bengo, Mr Revill, Dr Nkhoma and Mr Ng’ambi are supported by UK Research and Innovation as part of the Global Challenges Research Fund, grant number MR/P028004/1

TM, CW, PW, OJW, AG and TBH acknowledge joint Centre funding from the UK Medical Research Council and Department for International Development. Grant reference: MR/R015600/1

We would like to thank the Imperial College COVID-19 Response Team for their feedback and assistance with method development. We would also like to thank the University of Malawi Health Economics and Policy Unit Think Tank members for their support and collaboration in this project.

## Notes

### Competing Interest Statement

Prof Hallett reports grants from MRC, during the conduct of the study; grants from BMGF, MRC, UNAIDS, WHO, NIH, personal fees from The Global Fund, WHO, BMGF, outside the submitted work;.

### Author Declarations

No IRB/ethical approval was required as this is a theoretical modelling study using publicly available data

