## Supplementary Information for "The potential impact of intervention strategies on COVID-19 transmission in Malawi: A mathematical modelling study"

Estimates of disease severity

Following Walker *et al*., we distinguish three levels of disease severity for COVID-19: (i) those that do not require hospitalisation; (ii) those that do require hospitalisation but not intensive care; (iii) those that require intensive care (ICU). Two parameters govern the proportions in each category: the proportion of infected cases requiring hospitalisation (*p_severe*) and the proportion of hospitalised cases requiring ICU (*p_critical*).

Uncertainty in these parameters is captured using prior information reported from high-income countries and generating sets of probabilities conditional on the resulting IFR falling within the bounds described in the methods. To do this, we sampled from a range of possible values for each parameter (detailed in Supplementary Table 5) based on values observed in high-income settings, keeping the age-distribution consistent with that reported in Verity *et al* and Walker *et al*.[1, 2] As we have based the calculation of IFR so far on data from China, we assumed that all persons that needed hospitalisation or intensive care received it, and that the proportion of those requiring intensive care that die, *m*, is 50%. This is a generalised assumption which incorporates both the availability of treatment and the mortality rates across both the untreated and treated critical cases. We assume no mortality in severe cases in this instance, although this assumption is changed in the simulation model. Combining the assumptions together, gives an overall IFR across the 17 age-groups as follows:

${IFR}_{h}=\sum_{a=1}^{17} \left( {p\_severe}_{h,a}*{p\_critical}_{h,a}*m \right)*N_{a}$

Parameter sets for *p_severe* and *p_critical* were accepted if the resulting overall IFR fell within the 95% uncertainty interval of the adjusted IFR for Malawi and the process was repeated until 1000 accepted parameter sets were generated. The resulting parameter sets were used as inputs to the epidemic model.

Supplementary Table 1. Prevalence data used for the calculations of disease severity and infection fatality ratios.

| **Data** | **Source** | **Notes** |
| --- | --- | --- |
| TB | WHO TB Report  [3] | The estimated incidence of active TB in 2018 is used, which is the latest year for which data were available: number of active cases divided by population size for every age-group and by sex |
| HIV | UNAIDS Country Projections 2019  [4]  IHME  [5] | For Malawi: includes all PLHIV, irrespective of treatment status  For China: includes all PLHIV, irrespective of treatment status |
| Malaria | IHME  [5] | For Malawi only. Prevalence of clinical malaria episodes in 2017 was used, which is the latest year for which data were available |
| COPD | IHME  [5] | For both countries |
| CVD | IHME  [5] | For both countries  Included as part of the CVD comorbidities |
| CKD | IHME  [5] | For both countries  Included as part of the CVD comorbidities |
| Diabetes | Price *et al* [6]  IHME [5] | Included as part of the CVD comorbidities  Price et al 2018 for Malawi estimates and IHME 2020 for China estimates |
| Hypertension | Price *et al* [6] | Included as part of the CVD comorbidities  Price et al 2018 for Malawi estimates. Hypertension is included in the CVD prevalence estimates for the China |
| Obesity | Global Obesity Observatory  [7] | Included as part of the CVD comorbidities  For both countries |
| Malnutrition | IHME  [5] | For both countries |
| Population size | UNdata  [8] | For both countries |

All disease prevalence estimates are age- and sex-stratified.

Supplementary Table 2. Parameters used in the ICL Global COVID-19 Model

| Parameter | Value | Notes / source |
| --- | --- | --- |
| *R_0_* | 2 | range 1.5-3.0 |
| Hospital bed capacity | 1.3 beds per 1,000 population | Equates to 24,869 hospital beds. Source: World Bank [9] |
| ICU bed capacity | 25 |  |
| Mean latent period | 4.6 days | Estimated at 5.1 days. The last 0.5 days are incorporated in the infectious periods to capture pre-symptomatic infectivity |
| Mean duration of mild infection | 2.1 days | Incorporates 0.5 days of infectiousness prior to symptoms. In combination with mean duration of severe illness this gives a mean serial interval of 6.75 days |
| Mean duration of severe infection prior to hospitalisation | 4.5 days | Mean onset-to-admission of 4 days based on unpublished analysis of data from the ICNARC study. Includes 0.5 days of infectiousness prior to symptom onset |
| Mean duration of hospitalisation for non-critical cases if survive | 9.5 days | Based on unpublished analysis of data from the ICNARC study |
| Mean duration of hospitalisation for non-critical cases if die | 7.6 days | Based on unpublished analysis of data from the ICNARC study |
| Mean duration in ICU if survive | 11.3 days | Based on data from the ICNARC study adjusted for censoring |
| Mean duration in ICU if die | 10.1 days | Based on data from the ICNARC study adjusted for censoring. |
| Mean duration in recovery after ICU | 3.4 days | Based on unpublished analysis of data from the ICNARC study |
| Relative risk of mortality for patients with HIV | 2.75 | 95% CI 2.09 – 3.61 [10] |
| Relative risk of mortality for patients with TB | 2.58 | 95% CI 1.53 – 4.37 [10] |
| Relative risk of mortality for patients with malaria | 1.5 | Assumption, range 1-3 |
| Relative risk of mortality for patients with cardiovascular disease | 2.35 | 95% CI 1.44 – 3.84 [11] |
| Relative risk of mortality for patients with COPD | 1.76 | 95% CI 0.92 – 3.36 [11] |
| Relative risk of mortality for patients with obesity | 1 | No additional risk once controlling for other comorbidities [12] |
| Relative risk of mortality for patients with malnutrition | 1 | Assumption, range 1-3 |

Supplementary Table 3. Age-dependent mortality rates

|  | Severe cases |  | Critical cases |  |
| --- | --- | --- | --- | --- |
| Age-group | Probability death  with treatment | Probability death  no treatment | Probability death  with treatment | Probability death  no treatment |
| 0-4 | 0.013 | 0.026 | 0.5 | 0.95 |
| 5-9 | 0.013 | 0.026 | 0.5 | 0.95 |
| 10-14 | 0.013 | 0.026 | 0.5 | 0.95 |
| 15-19 | 0.013 | 0.026 | 0.5 | 0.95 |
| 20-24 | 0.013 | 0.026 | 0.5 | 0.95 |
| 25-29 | 0.013 | 0.026 | 0.5 | 0.95 |
| 30-34 | 0.013 | 0.026 | 0.5 | 0.95 |
| 35-39 | 0.013 | 0.026 | 0.5 | 0.95 |
| 40-44 | 0.015 | 0.026 | 0.5 | 0.95 |
| 45-49 | 0.019 | 0.026 | 0.5 | 0.95 |
| 50-54 | 0.027 | 0.026 | 0.5 | 0.95 |
| 55-59 | 0.042 | 0.084 | 0.5 | 0.95 |
| 60-64 | 0.069 | 0.138 | 0.5 | 0.95 |
| 65-69 | 0.105 | 0.210 | 0.5 | 0.95 |
| 70-74 | 0.149 | 0.298 | 0.5 | 0.95 |
| 75-79 | 0.203 | 0.406 | 0.5 | 0.95 |
| 80+ | 0.580 | 1.000 | 0.5 | 0.95 |

The probability of death in severe cases not receiving treatment has been changed from the default values in the ICL Global Model. Here we assume mortality rates for untreated severe cases are double those of the treated cases. Values are derived using data from the ICNARC study in the UK.[13]

Supplementary Table 4. Social contact matrix between age-groups in Zimbabwe

| Age-group | 0-4 | 5-9 | 10-14 | 15-19 | 20-24 | 25-29 | 30-34 | 35-39 | 40-44 | 45-49 | 50-54 | 55-59 | 60-64 | 65-69 | 70-74 | 75+ |
| --- | --- | --- | --- | --- | --- | --- | --- | --- | --- | --- | --- | --- | --- | --- | --- | --- |
| 0-4 | 1.006 | 1.030 | 0.883 | 0.732 | 0.647 | 0.712 | 0.645 | 0.463 | 0.289 | 0.214 | 0.211 | 0.215 | 0.179 | 0.121 | 0.075 | 0.048 |
| 5-9 | 0.807 | 2.863 | 2.668 | 0.968 | 0.423 | 0.449 | 0.468 | 0.416 | 0.289 | 0.212 | 0.182 | 0.186 | 0.177 | 0.140 | 0.093 | 0.066 |
| 10-14 | 0.659 | 2.544 | 3.766 | 1.865 | 0.580 | 0.413 | 0.452 | 0.536 | 0.375 | 0.251 | 0.207 | 0.208 | 0.193 | 0.177 | 0.136 | 0.096 |
| 15-19 | 0.617 | 1.041 | 2.104 | 2.548 | 1.070 | 0.575 | 0.536 | 0.645 | 0.441 | 0.297 | 0.267 | 0.264 | 0.229 | 0.225 | 0.182 | 0.112 |
| 20-24 | 0.947 | 0.790 | 1.135 | 1.858 | 1.363 | 0.969 | 0.752 | 0.723 | 0.474 | 0.363 | 0.333 | 0.310 | 0.281 | 0.254 | 0.199 | 0.123 |
| 25-29 | 1.091 | 0.878 | 0.848 | 1.046 | 1.014 | 1.131 | 0.885 | 0.707 | 0.425 | 0.330 | 0.297 | 0.299 | 0.271 | 0.225 | 0.153 | 0.101 |
| 30-34 | 1.268 | 1.175 | 1.191 | 1.252 | 1.010 | 1.136 | 1.119 | 1.032 | 0.593 | 0.405 | 0.356 | 0.364 | 0.341 | 0.249 | 0.164 | 0.102 |
| 35-39 | 0.976 | 1.119 | 1.512 | 1.612 | 1.041 | 0.972 | 1.105 | 1.221 | 0.756 | 0.485 | 0.420 | 0.409 | 0.358 | 0.276 | 0.177 | 0.105 |
| 40-44 | 0.960 | 1.225 | 1.668 | 1.738 | 1.076 | 0.923 | 1.001 | 1.192 | 0.863 | 0.629 | 0.488 | 0.434 | 0.378 | 0.311 | 0.207 | 0.117 |
| 45-49 | 0.796 | 1.007 | 1.248 | 1.312 | 0.922 | 0.801 | 0.766 | 0.856 | 0.704 | 0.549 | 0.446 | 0.383 | 0.352 | 0.298 | 0.215 | 0.120 |
| 50-54 | 0.844 | 0.928 | 1.106 | 1.266 | 0.909 | 0.774 | 0.722 | 0.796 | 0.587 | 0.479 | 0.417 | 0.407 | 0.397 | 0.331 | 0.242 | 0.159 |
| 55-59 | 0.902 | 0.993 | 1.163 | 1.312 | 0.888 | 0.818 | 0.776 | 0.812 | 0.547 | 0.431 | 0.426 | 0.483 | 0.484 | 0.382 | 0.281 | 0.210 |
| 60-64 | 0.823 | 1.041 | 1.191 | 1.250 | 0.882 | 0.815 | 0.797 | 0.781 | 0.524 | 0.435 | 0.457 | 0.532 | 0.567 | 0.449 | 0.347 | 0.259 |
| 65-69 | 0.695 | 1.026 | 1.357 | 1.534 | 0.995 | 0.841 | 0.725 | 0.751 | 0.537 | 0.459 | 0.475 | 0.524 | 0.560 | 0.471 | 0.372 | 0.333 |
| 70-74 | 0.592 | 0.943 | 1.442 | 1.712 | 1.080 | 0.791 | 0.664 | 0.668 | 0.495 | 0.458 | 0.481 | 0.532 | 0.599 | 0.515 | 0.455 | 0.383 |
| 75+ | 0.557 | 0.974 | 1.485 | 1.530 | 0.967 | 0.763 | 0.597 | 0.577 | 0.406 | 0.373 | 0.458 | 0.579 | 0.648 | 0.669 | 0.556 | 0.400 |

Values are estimated number of contacts between persons in each age-group per day. The contact matrix is adjusted to give age-specific contact rates.[14]

Supplementary Table 4. List of the interventions under consideration along with their implementation in the model.

| **Strategy** | **Implementation** |
| --- | --- |
| Unmitigated epidemic baseline | Includes only isolation of hospitalised cases |
| **Current situation:**  Implemented from 18^th^ April 2020 | Consider these strategies as a bundle equating to a combined reduction in *Rt* of 7% and a reduction in contact rates for the whole population plus additional reductions for employed people and school-aged children as detailed below.  Dates and details of individual non-pharmaceutical interventions are reported by OxCGRT[15] |
| Social distancing recommended | Reduce contact rates for the whole population by 24% (maximum reduction in population movement reported by Google Mobility Reports)[16]^#^ |
| Workplace closures | Reduce contact rates of those employed in offices and with access to internet at home (6·4% of the population aged 15-49) by 50% (this value will include social distancing so additional reductions of 26% are applied to the eligible population)[17] |
| School closures | Reduce contact rates between children aged 5-15 years by 50% (this value will include social distancing so additional reductions of 26% are applied to the eligible population) |
| Public events cancelled | Relative reduction in *Rt* 4% range 1-17%. We take the mid-point of the estimated relative reduction in *Rt* for each of these individual strategies[18] |
| Self-quarantine for returning individuals | Relative reduction in *Rt* 3% range 0-15%. We take the mid-point of the estimated relative reduction in *Rt* for each of these individual strategies[18] |
| Restriction of gatherings to 10 | Assume no impact above that of cancelling public events |
| International travel controls | Assume no new infections imported from other countries |
| **Enhanced Shielding of the Elderly:**  Current situation, plus:  Shielding the elderly | Reduce contact rates by 40% for populations aged ≥60 years in addition to the reduction in *Rt* and contact rates above.  It is implemented after a trigger is reached^$^ |
| **Lockdown:**  Current situation, plus:  Shelter in place  Restrict movements to essential activities such as the provision of essential goods and services  Enforce social distancing in excepted businesses  Prohibit public transportation  Prohibit all gatherings outside household | Consider that this bundle equates to a sustained reduction in *Rt* to 1. Estimated Rt <1 has been reported in low-income settings following lockdowns, we assume *Rt* would not fall below 1 due to continued household transmission and potential non-compliance.[19] The reductions in contact rates listed under “current situation” are also assumed to apply in this scenario.  It is implemented after a trigger is reached^$^ |

^#^Data are from Zambia, data from Malawi are not available. Note Zambia rural population is 56·5% compared with Malawi 83·1%.

^$^The trigger date for interventions to be applied was 1·0 COVID-19 deaths per 100,000 population per week

Supplementary Table 6. Estimates of disease severity for the Malawian population

| **Age-group**  **(years)** | **Proportion cases requiring hospitalisation** | | **Proportion hospitalised cases requiring ICU** | |
| --- | --- | --- | --- | --- |
|  | Credible range | Median accepted values  (2.5^th^ and 97.5^th^ quantiles) * | Credible range | Median accepted values  (2.5^th^ and 97.5^th^ quantiles) * |
| 0-4 | 0.0002-0.005 | 0.0013  (0.0003 - 0.0028) | 0.01-0.25 | 0.0624  (0.016 - 0.1434) |
| 5-9 | 0.0002-0.005 | 0.0013  (0.0003 - 0.0028) | 0.01-0.25 | 0.0624  (0.016 - 0.1434) |
| 10-14 | 0.0002-0.005 | 0.0013  (0.0003 - 0.0028) | 0.01-0.25 | 0.0624  (0.016 - 0.1434) |
| 15-19 | 0.0004-0.01 | 0.0026  (0.0007 - 0.0057) | 0.01-0.25 | 0.0624  (0.016 - 0.1434) |
| 20-24 | 0.001-0.025 | 0.0064  (0.0017 - 0.0142) | 0.01-0.25 | 0.0624  (0.016 - 0.1434) |
| 25-29 | 0.002-0.05 | 0.0128  (0.0034 - 0.0284) | 0.01-0.25 | 0.0624  (0.016 - 0.1434) |
| 30-34 | 0.0032-0.08 | 0.0205  (0.0054 - 0.0454) | 0.01-0.25 | 0.0624  (0.016 - 0.1434) |
| 35-39 | 0.0046-0.115 | 0.0295  (0.0077 - 0.0653) | 0.0106-0.265 | 0.0662  (0.0169 - 0.152) |
| 40-44 | 0.0058-0.145 | 0.0372  (0.0097 - 0.0823) | 0.012-0.3 | 0.0749  (0.0192 - 0.1721) |
| 45-49 | 0.0078-0.195 | 0.05  (0.0131 - 0.1107) | 0.015-0.375 | 0.0937  (0.024 - 0.2151) |
| 50-54 | 0.0116-0.29 | 0.0744  (0.0195 - 0.1646) | 0.0208-0.52 | 0.1299  (0.0332 - 0.2983) |
| 55-59 | 0.0144-0.36 | 0.0923  (0.0242 - 0.2044) | 0.0298-0.745 | 0.1861  (0.0476 - 0.4274) |
| 60-64 | 0.0204-0.51 | 0.1308  (0.0342 - 0.2895) | 0.0448-1.12 | 0.2797  (0.0716 - 0.6425) |
| 65-69 | 0.0234-0.585 | 0.15  (0.0393 - 0.3321) | 0.0614-1.535 | 0.3834  (0.0981 - 0.8806) |
| 70-74 | 0.0292-0.73 | 0.1872  (0.049 - 0.4144) | 0.0772-1.93 | 0.4821  (0.1233 - 1) |
| 75-79 | 0.0354-0.885 | 0.2269  (0.0594 - 0.5024) | 0.0922-2.305 | 0.5757  (0.1473 - 1) |
| 80+ | 0.036-0.9 | 0.2308  (0.0604 - 0.5109) | 0.1418-3.545 | 0.8854  (0.2265 - 1) |

* distributions obtained through rejection sampling.

Supplementary Table 7. Simulation outputs under different assumptions of *R_0_*.

|  |  | Baseline | Current situation | Enhanced shielding | Lockdown |
| --- | --- | --- | --- | --- | --- |
| *R_0_* = 1.5 | Total cases / 1,000 | 553  (462-642) | 319  (258-370) | 163  (130-195) | 40  (27-52) |
|  | Number hospital beds required at peak | 23300  (13,400-33,500) | 16800  (9,500-27,300) | 7600  (4,600-12,100) | 1200  (700-2,300) |
|  | Number ICU beds required at peak | 1,000  (600-1,500) | 700  (500-1,100) | 200  (200-300) | 100  (100-100) |
|  | Total number of deaths | 56900  (36,100-83,800) | 39800  (25,600-60,100) | 11500  (7,800-17,200) | 5400  (3,200-8,400) |
| *R_0_* = 2.0 | Total cases / 1,000 | 770  (666-875) | 584  (507-662) | 513  (436-595) | 71  (58-85) |
|  | Number hospital beds required at peak | 40,000  (27,800-59,900) | 35,600  (25,300-53,100) | 27,900  (17,800-38,000) | 2,100  (1,100-3,500) |
|  | Number ICU beds required at peak | 2,100  (1,400-3,400) | 1,800  (1,200-2,900) | 900  (600-1,400) | 100  (100-200) |
|  | Total number of deaths | 80,400  (49,500-118,800) | 77,200  (48,000-114,300) | 46,700  (30,400-68,700) | 10,100  (6,500-15,300) |
| *R_0_* = 3.0 | Total cases / 1,000 | 920  (815-1,026) | 756  (669-844) | 716  (627-808) | 109  (93-125) |
|  | Number hospital beds required at peak | 61,700  (41,200-98,800) | 57,600  (38,700-92,000) | 49,700  (34,200-75,400) | 4,100  (2,400-6,900) |
|  | Number ICU beds required at peak | 3,800  (2,500-6,000) | 3,500  (2,300-5,600) | 2,300  (1,500-3,700) | 200  (100-300) |
|  | Total number of deaths | 93,000  (57,700-136,400) | 92,100  (57,200-134,800) | 72,000  (44,900-105,500) | 15,800  (10,400-23,600) |

All values are medians of 1000 simulations using the sampled parameter sets. Numbers of hospital beds, ICU beds and deaths are rounded to the nearest 100. The trigger day for interventions (1.0 death per 100,000 population per week) is day 145, 82, and 52 for *R_0_*=1.5, 2.0 and 3.0 respectively.

Supplementary Table 8. Projected cumulative number of deaths with increases in hospital capacity.

|  | Low mortality | High mortality |
| --- | --- | --- |
| Intervention | Total deaths per 1,000 population | Total deaths per 1,000 population |
| Current capacity | 4·04  (2·51-5·98) | 5·96  (2·99-10·62) |
| Increase capacity by 25% | 3·95  (2·48-5·89) | 5·41  (2·75-9·75) |
| Increase capacity by 50% | 3·89  (2·47-5·79) | 4·94  (2·58-8·98) |
| Increase capacity by 75% | 3·82  (2·47-5·70) | 4·66  (2·51-8·43) |
| Increase capacity by 100% | 3·79  (2·45-5·61) | 4·41  (2·48-7·87) |

Low and high mortality refer to the assumptions on mortality rates of untreated severe cases. Low mortality rates are those used in the main analysis (detailed in Supplementary Table 3) and high mortality applies a mortality rate of 0·6 across all age-groups.

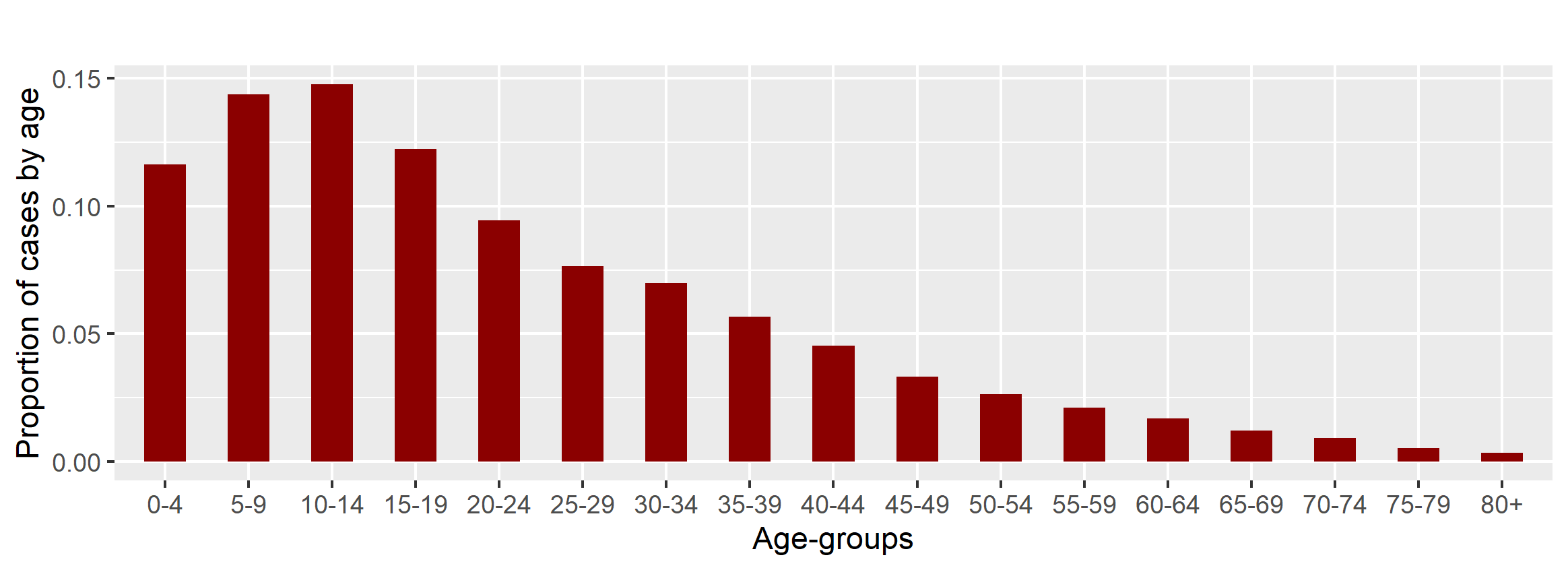

Supplementary Figure 1. Age distribution of infected cases (asymptomatic and symptomatic) at the peak of the projected unmitigated epidemic.

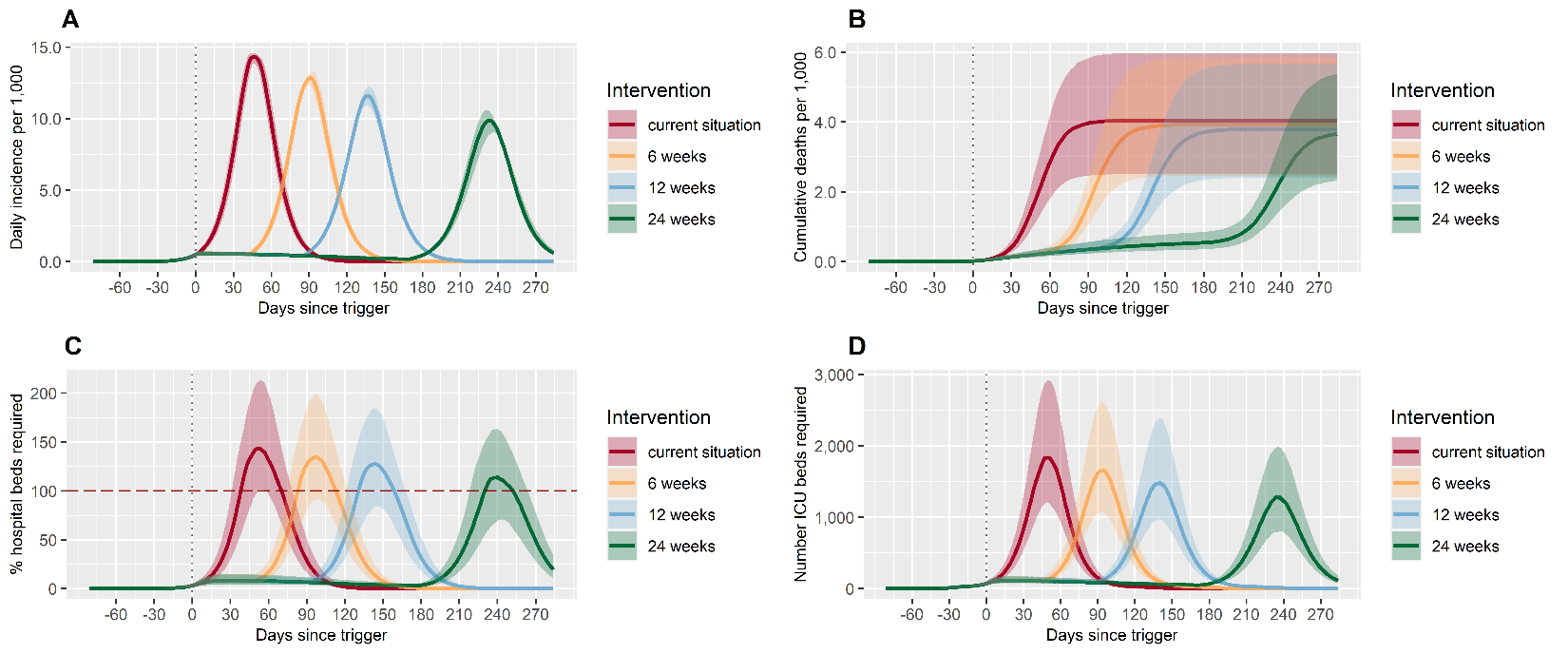

Supplementary Figure 2. Impact of changing the duration of lockdown compared with the current situation on the daily incidence per 1,000 population (A), the cumulative deaths per 1,000 population (B), the percentage of hospital beds that are required (C) and the number of ICU beds that are required (D). The current situation reflects the non-pharmaceutical interventions adopted by Malawi at the beginning of the outbreak. Each intervention represents the implementation of lockdown over 6, 12 or 24 weeks. The trigger date (1 death per 1,000 population per week) is shown with a vertical grey dashed line. The red horizontal dashed line shows the capacity of the health system for non-intensive care (C).

3.	World Health Organization, *WHO TB burden estimates*. 2020: Geneva, Switzerland.

4.	UNAIDS, *AIDSInfo*. 2020: Geneva, Switzerland.

5.	Global Burden of Disease Collaborative Network, *Global Burden of Disease Study 2017 (GBD 2017) Results*. 2018, Institute for Health Metrics and Evaluation (IHME): Seattle, USA.

6.	Price, A.J., et al., *Prevalence of obesity, hypertension, and diabetes, and cascade of care in sub-Saharan Africa: a cross-sectional, population-based study in rural and urban Malawi.* The lancet Diabetes & endocrinology, 2018. **6**(3): p. 208-222.

7.	World Obesity Federation, *Global Obesity Observatory*. 2020.

8.	United Nations Statistics Division, *Population by age, sex and urban/rural residence*. 2020: New York, USA.

9.	The World Bank, *World Bank Open Data [Hospital beds per 1,000 people]*. 2020.

10.	Davies, M.-A. and Western Cape Department of Health. *Western Cape: COVID-19 and HIV / Tuberculosis*. 2020 10th June 2020]; Available from: <https://storage.googleapis.com/stateless-bhekisisa-website/wordpress-uploads/2020/06/df4fcf94-covid_update_bhekisisa_wc_3.pdf>.

11.	Ssentongo, P., et al., *The association of cardiovascular disease and other pre-existing comorbidities with COVID-19 mortality: A systematic review and meta-analysis.* medRxiv, 2020.

12.	Palaiodimos, L., et al., *Severe obesity, increasing age and male sex are independently associated with worse in-hospital outcomes, and higher in-hospital mortality, in a cohort of patients with COVID-19 in the Bronx, New York.* Metabolism, 2020. **108**: p. 154262.

13.	Intensive Care National Audit & Research Centre *ICNARC report of COVID-19 in critical care*. 2020.

14.	Melegaro, A., et al., *Social contact structures and time use patterns in the Manicaland Province of Zimbabwe.* PloS one, 2017. **12**(1).

15.	Hale, T., et al., *Oxford COVID-19 Government Response Tracker, Blavatnik School of Government*. 2020: University of Oxford.

16.	Google LLC, *Google COVID-19 Community Mobility Reports*. 2020.

17.	National Statistical Office and ICF, *Malawi Demographic and Health Survey 2015–2016.* 2017.

18.	Flaxman, S., et al., *Estimating the effects of non-pharmaceutical interventions on COVID-19 in Europe.* Nature, 2020.

19.	MRC Centre for Global Infectious Disease Analysis, I.C.L., *Situation Report for COVID-19: Africa, 2020-08-17*. 2020, Imperial College London: London, UK.
